# On discrete time epidemic models in Kermack-McKendrick form

**DOI:** 10.1101/2021.03.26.21254385

**Authors:** Odo Diekmann, Hans G. Othmer, Robert Planqué, Martin C. J. Bootsma

**Affiliations:** Mathematical Institute, Utrecht University, Utrecht, The Netherlands; School of Mathematics, University of Minnesota, Minneapolis, USA; Department of Mathematics, Vrije Universiteit Amsterdam, Amsterdam, The Netherlands; Department of Epidemiology, Universitair Medisch Centrum Utrecht, Utrecht, The Netherlands

## Abstract

Surprisingly, the discrete-time version of the general 1927 Kermack-McKendrick epidemic model has, to our knowledge, not been formulated in the literature, and we rectify this omission here. The discrete time version is as general and flexible as its continuous-time counterpart, and contains numerous compartmental models as special cases. In contrast to the continuous time version, the discrete time version of the model is very easy to implement computationally, and thus promises to become a powerful tool for exploring control scenarios for specific infectious diseases. To demonstrate the potential, we investigate numerically how the incidence-peak size depends on model ingredients. We find that, with the same reproduction number and initial speed of epidemic spread, compartmental models systematically predict lower peak sizes than models that use a fixed duration for the latent and infectious periods.

## 1 Introduction

The day-night cycle has a strong impact on the behaviour of humans, animals and plants. As a rule, the resulting time heterogeneity is ignored in epidemiological and ecological models. One simply pretends that the representation of time by a continuous quantity *t*, “flowing” at a constant rate, is suitable for bookkeeping of the time course of the relevant events.

Census data, on the other hand, are often collected at regular intervals, so on a discrete time basis. Indeed, as evidenced by the Covid-19 pandemic, incidence is usually reported in the form of the number of new cases on a particular day or in a specified week.

So even when the processes that we want to capture take place in continuous time, one may want to consider discrete time bookkeeping schemes, in order to relate directly to the data, as advocated in the pioneering paper [2]. Moreover, a significant bonus of discrete time models is that numerical implementation is straightforward and that, accordingly, simulations are easy to perform. In sharp contrast, the numerical solution of continuous time renewal equations, as discussed in [3], presents a substantial challenge to the uninitiated.

Counter to the practical advantages runs a modelling difficulty: the formulation of discrete time models is subtle and hence error-prone. In infinitesimal time intervals the effects of different mechanisms are independent. Consequently one can add terms that describe contributions to the rate of change of a quantity. When trying to capture (in one go, rather than by solving a differential equation) change in a finite time interval, we do have to think about the order of events and how one event may trigger or prevent another event. In the epidemic context a key point is that, when a susceptible host is infected by an infectious host, it cannot any longer be infected by another infectious host.

The first aim of this short note is to formulate the discrete time version of the general Kermack-McKendrick epidemic model from 1927. As far as we know, this has not been done before. And yet the model is, most likely, an ideal tool for data driven analysis of infectious disease outbreaks since, as we shall demonstrate, it is not only general and flexible, but also extremely user friendly. A second (and admittedly somewhat pedantic) aim of this note is to point out explicitly a frequent mistake in the formulation of discrete time epidemiological and ecological models. We hope that by clearly exposing the underlying fallacy, the mistake will have had its day.

To put some flesh on the bones, we show that the qualitative behaviour of the discrete time model is the spitting image of the qualitative behaviour of the continuous time model. In order not to ignore the popularity of compartmental variants (which is unwarranted, in our opinion, as there is neither evidence that the length of, for instance, the infectious period is exponentially/geometrically distributed nor that infectiousness is constant during this period), we put them in the spotlight. To conclude, we illustrate the relevance (in particular for public health policy) of the lesser known members of the Kermack-McKendrick family, by investigating numerically how the peak of the incidence curve varies among members that are identical with respect to both the initial growth rate *ρ* and the basic reproduction number *R*_0_, but differ in assumptions about the duration of the exposed and the infectious period.

To avoid misunderstanding, we now clarify what is, and what is not, stochastic in the models formulated and analyzed below. All models are deterministic at the population level. (One can think of them, in Kurtz spirit, see [16], as the large initial population size limit of a stochastic model for finitely many individuals.)

When one assumes that the infectious period of all individuals, once infected, has exactly the same length and their infectiousness during this period is one and the same constant, there is no randomness at the individual level either.

But most models incorporate heterogeneity/stochasticity at the individual level. For instance, in the familiar continuous time *SIR* compartmental model, the length of the infectious period of a newly infected individual is exponentially distributed, say with parameter *γ*. If during the infectious period all infected individuals have the same constant infectiousness *β*, then the expected infectiousness *A*(*τ*) at time *τ* after becoming infected equals *βe*^*−γτ*^. This reflects that after time *τ* has elapsed, the probability to be still infectious equals *e*^*−γτ*^. At the population level, we translate this into: a fraction *e*^*−γτ*^ of those infected at time *t* is still infectious at time *t* + *τ*. In the discrete time setting, we should replace the exponential distribution by the geometric distribution. As a final elucidation we mention that the parameter *β* and the function *A*(*τ*) implicitly incorporate information about the contact (between individuals) process that underlies transmission. A key feature is that contacts are assumed to be uniformly at random (so neither spatial nor age nor social structure are incorporated).

## 2 The cumulative force of infection

First of all we want to motivate and promote the equation

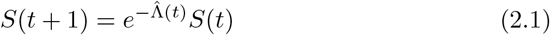

as a buiding block for discrete time models of the spread of an infectious disease in a host population when

- the disease generates permanent immunity,
- the host population is demographically closed (meaning that demographic turnover happens at a much slower time scale than transmission of the disease and is therefore ignored).

As usual, *S*(*t*) denotes the size of the subpopulation of susceptibles at census time *t*. Underlying (2.1) is a choice of the unit of time: it equals the length of the interval between one census and the next. So the magnitude of 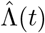 is proportional to this length and when this magnitude figures in our discussion below, one may interpret the statements in terms of the length of the discretization step. We call 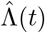 the cumulative force of infection in the time window (*t, t* + 1] for reasons that we now explain. The continuous time version of (2.1) reads

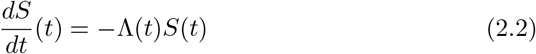

where Λ(*t*) is the force of infection at time *t*, i.e., the probability per unit of time for a susceptible to become infected at time *t*. By integration we deduce from (2.2) the relation

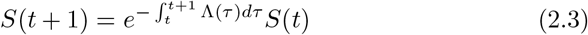

The first factor at the right hand side of (2.1) and (2.3) is, in both cases, the probability for a susceptible to escape from infection in the time window (*t, t*+1]. The integral

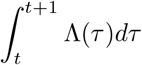

in (2.3) is replaced by 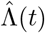 in (2.1) and this, we hope, clarifies why we call 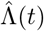 the cumulative force of infection.

We insist that one should adjust the multiplicative factor (as was indeed done in [2]) and *not* replace the differential equation (2.2) by the “additive” difference equation

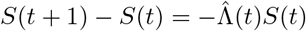

i.e., by

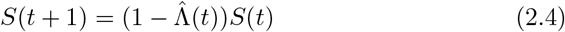

Of course (2.4) provides a good approximation of the “true” equation (2.1) for small values of 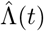. But (2.4) is not exact and it may fail dramatically for not so small values of 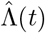, in particular since it may lead to negative values of *S*. The reason is that (2.4) does not take into account that a host can become infected only once. To stress this point, we now present a somewhat mechanistic derivation of the multiplicative factor in (2.1), showing that (2.1) does take this into account.

Assume that when a single infectious individual is present in a certain host population (during a time interval of, say, length one) every susceptible host becomes infected with probability *p*. Then any susceptible escapes from becoming infected with the complementary probability 1 *− p*. Next assume that there are *N* infectious individuals and that these make contacts with susceptibles independently of each other. Then any susceptible escapes from becoming infected with probability

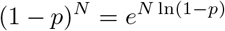

So a susceptible is infected with probability 1 *−* (1*− p*)^*N*^ rather than with “probability” *pN*.

From a numerical point of view, the exponential has the disadvantage of being expensive in terms of calculation costs. It may therefore be tempting to reduce the step size in order to work safely with the linear approximation. We actually wonder whether solving the relevant ODEs with for instance a Runge Kutta solver is not a more attractive alternative, especially in terms of accuracy. Moreover, we maintain that choosing a time interval that matches the data points has definite advantages.

## 3 The general discrete time Kermack-McKendrick model

As already expressed in (2.2), the incidence at time *t* equals Λ(*t*) *S*(*t*) with Λ the force of infection. Common sense tells us that the current force of infection is generated by individuals who were themselves infected some time ago. Following earlier work by Ross and Hudson (see [10, 11] and references in there), Kermack and McKendrick translate this observation into the constitutive equation

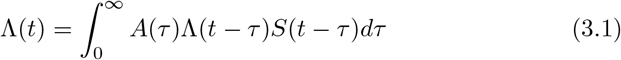

with *A*(*τ*) the *expected* contribution to the force of infection at time *τ* after infection. So in this top-down approach the infinite dimensional parameter *A* is introduced as a key model ingredient. For any specific disease one may, in principle, use a within-host model of the struggle between pathogen and immune system to provide bottom-up a quantitative specification. Alternatively, one may use population level data to infer (certain characteristics of) *A*, cf. [19]. Often this is done after first restricting to a parameterized family of functions *A* (but see [9] for an alternative methodology.) Note that, with *N* denoting the population size, we have

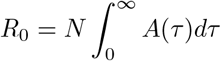

and that, in the initial phase of the epidemic, the distribution of the generation-interval (see [17, 18], and the references given in there) has as density the renormalized (to have integral 1) function *A*.

The incidence at time *t* is given by Λ(*t*)*S*(*t*). Under the “permanent immunity and no demographic turnover” assumption, the incidence equals *−S*^*′*^(*t*), cf. (2.2). Substituting this into (3.1), we deduce by integration (and upon changing the order of integration) the identity

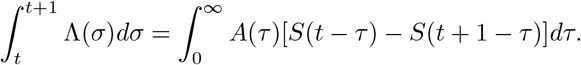

The discrete time counterpart reads

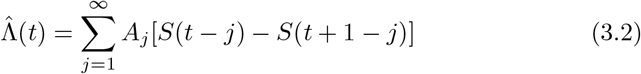

where now *A*_*j*_ is the expected contribution to the cumulative force of infection in (*t, t* + 1] of an individual who itself became infected in the time window (*t − j, t − j* + 1], so *j* time steps earlier, and *S*(*t − j*) *− S*(*t* + 1 *− j*) is the incidence in (*t − j, t − j* + 1]. So the key model ingredient is now the collection

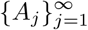

of positive/non-negative numbers, which we assume to be such that

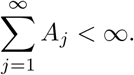

(Incidentally, note that control measures or seasonality may cause the *A*_*k*_ to depend on calendar time. See the end of Section 6 for a somewhat concrete example. In Section 7 we shall briefly indicate how one can easily implement this generalization.)

Equation (2.1), with 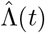 specified by (3.2), provides an updating scheme, but to get started one needs to specify an “initial” condition in the form of the history of *S* up to a certain point in time. The interpretation requires that this prescribed history is a monotone non-decreasing (when looking back into time) sequence, bounded from above by the total host population size. We shall denote this total size by *N*.

As we show next, one can reformulate (2.1), (3.2) as the scalar higher order recursion relation

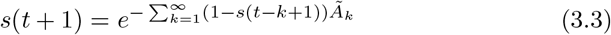

Where

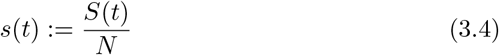

and

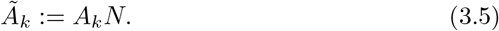

Equation (3.3) is the discrete time analogue of the nonlinear renewal equation

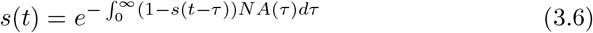

that follows by combining (2.2) with (3.2) and incidence equal to *−S*^*′*^, see [3, 6]. Both (3.3) and (3.6) involve the additional assumption

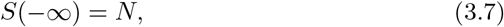

expressing that in the infinite past all host individuals were susceptible.

To derive (3.3), first note that iteration of (2.1) yields, if (3.7) holds, the identity

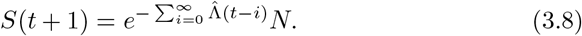

From (3.2) we deduce

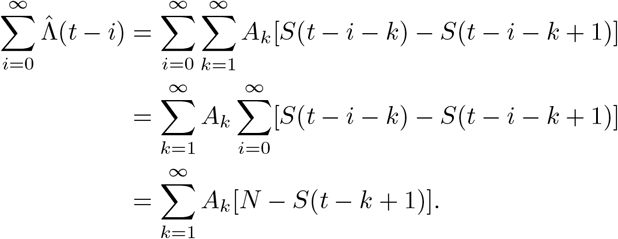

If we use this last identity in (3.8), divide both sides of (3.8) by *N* and adopt the notation (3.4) and (3.5), we obtain (3.3).

If one copies (3.3), with *t* + 1 replaced by *t*, and combines the two formulas, one can derive the variant

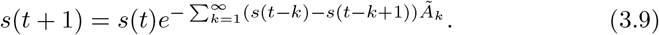

This variant has the advantage that one can provide an initial condition, say at time zero, by prescribing *s*(0) and the (nonnegative) incidences…, *s*(*−*3) *−s*(*−*2), *s*(*−*2) *−s*(*−* 1), *s*(*−*1) *−s*(0). We refer to Section 7 for a more pragmatic formulation of the initial value problem.

We conclude that (3.3)/(3.9) is the mathematical form of the discrete time Kermack-McKendrick model with, in principle, a countably infinite parameter 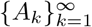, but in practice a finite dimensional parameter with an infinite tail of zeros.

## 4 The initial phase and the final size

To capture the demographic stochasticity during the very early phase of the introduction of an infectious disease in a host population, we need branching processes, see e.g. [6]. But once there is a large number of infected individuals, we can switch to a deterministic description. The large number may, of course, still constitute only a rather small *fraction* of a very large host population. In this situation we may put

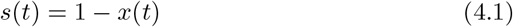

into (3.3) and assume that *x* is so small that it makes sense to replace the exponential by the zero’th and first order terms of its Taylor expansion. This yields the linearized equation

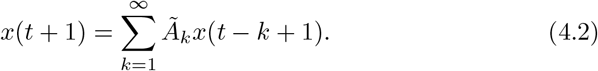

We define

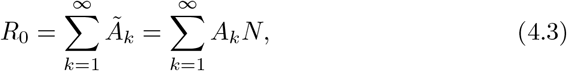

and interpret, based on the last identity, *R*_0_ as the expected number of secondary cases caused by a primary case in a totally susceptible host population.

In order to show that positive solutions of (4.2) grow when *R*_0_ *>* 1, but decline when *R*_0_ *<* 1, we make the Ansatz

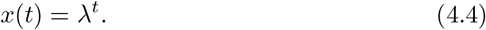

By substitution of (4.4) into (4.2) we find that *x* defined by (4.4) is indeed a solution if and only if *λ* is a real root of the discrete time characteristic equation

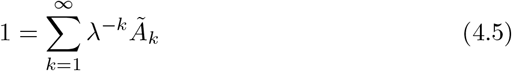

known as the Euler-Lotka equation. The non-negativity of *Ã* _*k*_, *k* = 1, 2,…, guarantees that (4.5) has at most one real root *ρ* and that it does indeed have a real root when the right hand side assumes a value bigger than one for some real *λ*, so in particular when *R*_0_ *>* 1 (when *R*_0_ *<* 1 and *Ã* _*k*_ has power-like behaviour for *k→ ∞*, the value of the right hand side may jump from a value less than one to infinity when *λ* is decreased; when *Ã* _*k*_ = 0 for large *k*, this cannot happen and *ρ* exists). Readers who wonder (or even worry) about the potential importance of complex roots can consult [6, Section 8.2] and the references given there, to be eased.

So we see that a key point is that the right hand side of (4.5) is a monotone decreasing function of real *λ*. And as a consequence we have

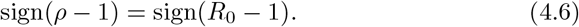

(Incidentally, note that *ρ* corresponds to *e*^*r*^, with *r* the Malthusian parameter featuring in the continuous time theory.) General linear theory, cf. [1], guarantees that positive solutions of (4.2) grow geometrically with rate *ρ* for *t→ ∞* when *ρ >* 1 (and decline with rate *ρ*, when *ρ* exists and is less than one). General nonlinear theory, cf. [20], guarantees that the steady state solution *s*(*t*)*≡* 1 of (3.3) is asymptotically stable for *ρ <* 1 (hence for *R*_0_ *<* 1), but unstable for *ρ >* 1, i.e., for *R*_0_ *>* 1 (here we refer to the Principle of Linearized Stability; the more general Hartman-Grobman Theorem implies that the intersection of the unstable manifold and the positive cone is one-dimensional; this means that, modulo translation, there is exactly one positive solution of (3.3) that has limit 1 for *t → −∞*, see [4] for the continuous time version).

So when *R*_0_ *>* 1, the introduction of the pathogen will, provided the pathogen does not go extinct by bad (or good, depending on the point of view) luck when still very rare, break through and cause *s* to decrease to below 1. The interpretation makes it obvious that *s* is a monotone decreasing function of time, and that it has a limit for *t → ∞*. We denote this limit by *s*(*∞*). The equation

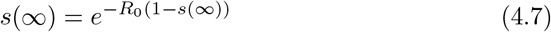

is obtained by passing to the limit in (3.3), while using that the {*Ã* _*k*_} are summable. For *R*_0_ *>* 1 this equation has a unique solution in (0, 1), see Figure 1, and [6, Exercise 1.19].

**Figure 1.**
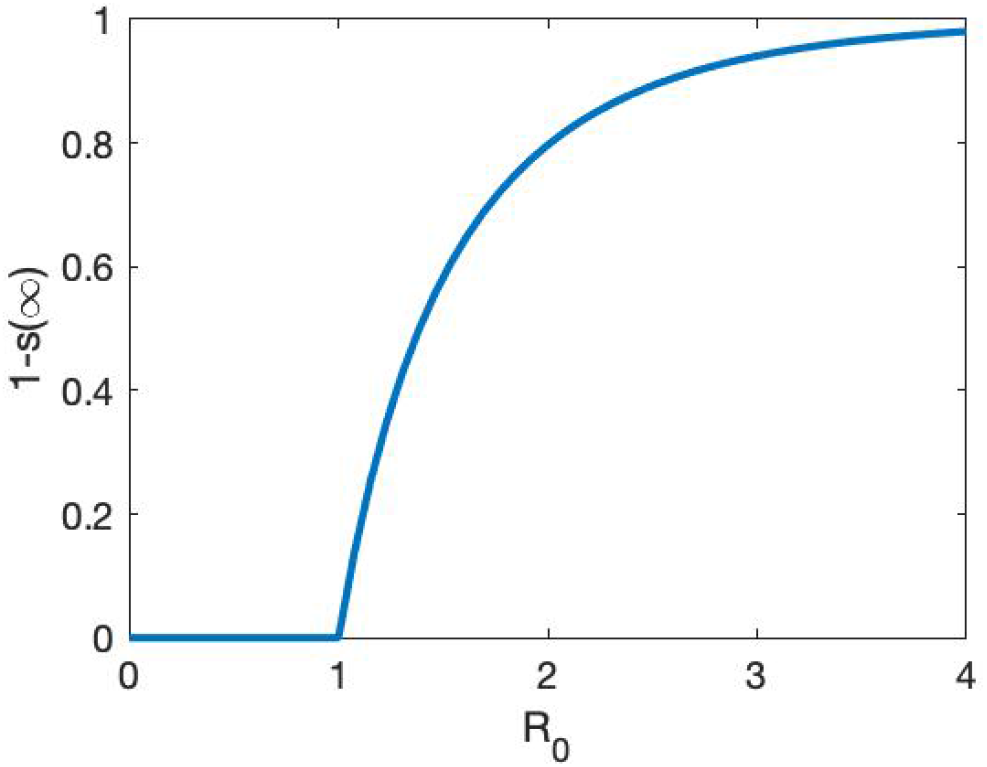
Graph of the final size 1 *− s*(*∞*), i.e., the fraction of the population that gets infected in the course of the outbreak, as a function of the basic reproduction number *R*_0_.

A comparison of the results in [13], [6, Chapter 1] and [3], with those above, establishes that when we compare the continuous time and discrete time formulations,

- there is only a formal difference in the expressions for *R*_0_;
- if we put *ρ* = *e*^*r*^, there is only a formal difference in the equations characterizing, respectively, *ρ* and *r*;
- the equations specifying *s*(*∞*) on the basis of *R*_0_ are identical (as already noted in [2]).

We conclude that at the level of theory, there is an exact parallel.

## 5 Compartmental formulation for some very special cases

We shall use the standard notational convention (or should one say “ambiguity”?) that a compartment and its contents are denoted by the same symbol. We start with *SIR* and after that generalize to *SEIR*, hoping that these two examples elucidate the general pattern how to construct discrete time models in compartmental settings. See [12] for a more general set-up.

Assume that, upon infection, an individual is transferred from the compartment *S* to the compartment *I* of infectious individuals. Assume that every following time step this infected individual stays in *I* with probability 1*− α* while being “removed” (i.e., losing infectiousness, either by way of the immune system conquering the pathogen, or by death) with probability *α*. We put removed individuals in a compartment *R* and assume that immunity is permanent (and resurrection impossible). Finally, we assume that the cumulative force of infection equals *βI*, i.e., the per capita contribution to the force of infection equals *β*. Note that *β* is proportional to the length of the discretization interval, i.e., the time between census points, and that 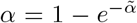 with 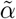 proportional to this length.

These assumptions lead to the system of recurrence relations

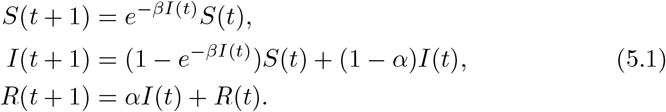

We show that the system (5.1) may be reduced to the scalar recurrence (3.3) by choosing the *Ã* _*k*_ appropriately:

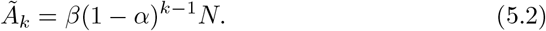

The first step corresponds to the derivation of (3.8): by iteration of the first equation of (5.1) we obtain

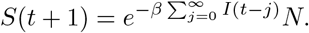

Rewriting the second equation of (5.1) as

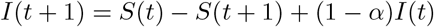

we obtain by summation the identity

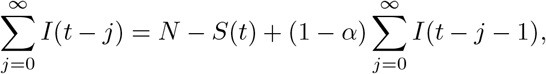

and by substitution of this identity repeatedly at the right hand side,

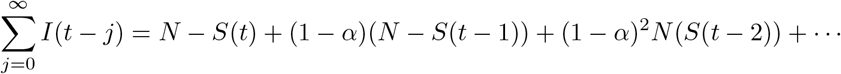

Finally, substitution of this last identity in the formula for *S*(*t* + 1) above yields (3.8).

Conversely, starting from (3.3) with *Ã* _*k*_ given by (5.2), we easily recover (5.1) by defining

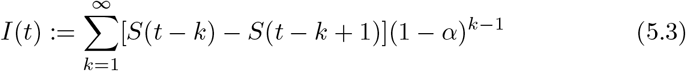

(note that the equation for *R*(*t*) is just an appendix; it has no impact on the dynamics of *S*(*t*) and *I*(*t*); it simply keeps track of individuals that are no longer infectious).

We emphasize that if one replaces *e*^*−βI*(*t*)^ by 1 *− βI*(*t*), the reduction to a higher order scalar recursion relation fails (we invite readers to convince themselves of this fact)!

In order to capture a latent period, we next change the assumptions. Upon infection, an individual now enters the compartment *E* of exposed (i.e., infected but not yet infectious) individuals. When the length of the latent period is geometrically distributed with parameter *γ*, we have to replace (5.1) by

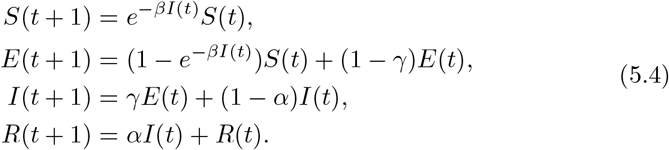

Do parameters *Ã* _*k*_ exist such that (5.4) can be condensed to (3.3)? It is helpful to think in terms of a stochastic process in which an individual can be in the states *S, E, I* and *R*. In fact, *E* and *I* suffice, since we start “looking” at the individual when it is infected and stop “looking” when it loses infectiousness. If we label *E* with index 1 and *I* with index 2, then the probability distribution of the state-at-infection is represented by the vector

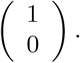

The state transitions are described by the matrix

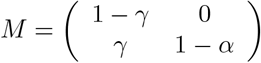

and infectiousness by the vector

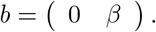

So the expected infectiousness *k* units of time after becoming infected is given by

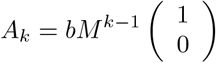

and hence by

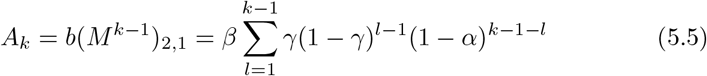

(with the convention that the sum equals zero when the upper index does not exceed or equal the lower index). The parameters *Ã* _*k*_ are again defined by (3.5). And when *Ã* _*k*_ has the form defined by (3.5) and (5.5), then (5.4) follows from (3.3) if we define

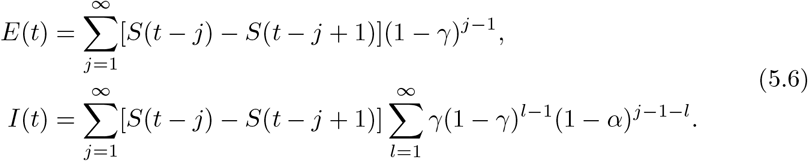

We trust that our presentation above, in terms of two vectors and one matrix, all having a well-defined interpretation, makes clear how one can in general relate compartmental epidemic models to a scalar higher order recursion. See Section 9.3 of [7] for a detailed elaboration of the continuous time case.

We conclude that there is a multitude of compartmental models that correspond to a special choice of the parameters *A*_*k*_. To prove the results of Section 4 directly for a model with 27 compartments is a hell of a job, especially if one does not recognize the underlying structure (and 27 components make recognition difficult). More importantly, it is an unnecessary job: one only needs to observe that one deals with a special case of (3.3).

## 6 On the choice of parameters *A*_*k*_

The ingredients {*A*_*k*_} subsume mechanistic properties of the process of contact between hosts as well as physiological/immunological properties of within-host dynamics. As a rule, information about such properties is scarce. One has to make educated guesses. See [15] for a concrete example.

The *SIR* and *SEIR* formulations of the last section have the advantage of involving just a few parameters. But, in our opinion, they have the disadvantage of being wrongly-educated guesses: they result from the tendency to do what others do, despite the fact that data, as a rule, do *not* support geometric distributions for the length of the latent- and/or the infectious period.

A facilitating aspect is that *A*_*k*_ are averages (see [6, Section 2.1] for a detailed exposition, including examples). If we “know” that at day six after infection only 10% of the infected individuals is infectious, while at day seven this rises to 20%, we can use this information directly in our choice of *A*_*k*_. If we know that at days six and seven the degree of infectiousness differs among individuals, we can still use the guestimated average.

A more theoretical example is the following. Assume that a fraction *p* of the infected individuals is asymptomatic. Assume that a symptomatic individual has at day *j* after infection a probability *θ*_*j*_ to be detected and next put into quarantine. Assume that the intrinsic infectiousness and contact intensity of symptomatic and asymptomatic cases is identical and given by {*B*_*k*_}. Then we choose

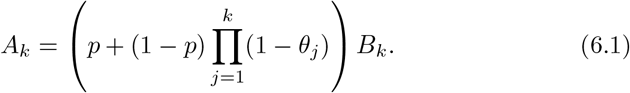

Note that (6.1) is based on the debatable assumption that at the day of its detection an individual does not contribute to the force of infection. This weakness is easily remedied, but at the cost of introducing yet another parameter.

The parameters *θ*_*j*_ can capture the effect of testing. During a serious outbreak, such as the Covid-19 pandemic, the testing policy and possibility depend on calendar day. This introduces time dependence in the parameters *θ*_*j*_. Similarly, control measures that reduce contact opportunities affect the *B*_*k*_ in a time-dependent multiplicative manner. In the next section we introduce a computational scheme in which such time dependence is easily incorporated.

## 7 Reformulation as a first order system

As Section 4 shows, the scalar higher order recursion relation (3.3) is very convenient for theoretical purposes. But for doing computations, a first order system of equations is more convenient.

For feasibility, we want a finite dimensional system. To achieve this, we make the very reasonable assumption that the indices *j* for which *A*_*j*_ is strictly positive have a finite upper bound. In other words, we assume that an integer *m* exists such that *A*_*j*_ = 0 for all *j ≥ m* + 1. The relevant consequence is that the history of *S*, that matters for determining the future, has finite length.

Define

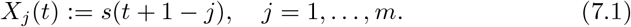

Much of the dynamics of the vector *X* amounts to shifting:

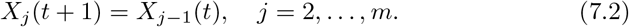

Combination of (2.1) and (3.3) yields the rule for extension

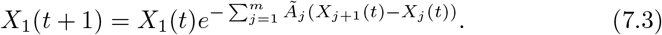

In (7.3) it is harmless to allow *Ã* _*j*_ to depend on time *t*!

Alternatively we might start from (3.9) and choose as before

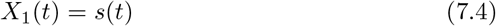

but for *j >* 1

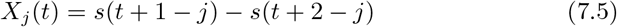

which corresponds to the incidence in time window *t* + 1*− j*. This leads to the update rules

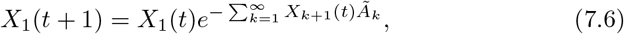

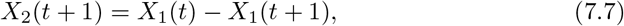

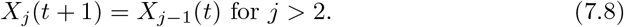

In this formulation too, we can allow *Ã* _*k*_ to depend on time *t*.

This seems a good moment to point out that the use of labels like ‘exposed’ or ‘infectious’ is perfectly possible within the general framework. For any such label, say *L*, specify, on the basis of the choice of the parameters *A*_*k*_ as described in Section 6, the probability *π*_*j*_ that an individual carries this label at time *j* after becoming infected. Then the number of individuals carrying label *L* at time *t* is given by

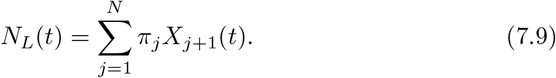

So all one needs to do to plot the time course of *N*_*L*_, is to add to (7.6)-(7.8) the equation (7.9) (with *t* replaced by *t* + 1, for consistency).

Note that (5.3) and (5.6) are examples of (7.9). In the very special situation considered in Section 5, the labels actually correspond to states at the individual level and as a consequence one can express *N*_*L*_(*t* + 1), for *L* = *S, E, I, R* in terms of these same quantities at time *t*, without reference to *X*(*t*). In general this is impossible. (Incidentally, note that probabilists often speak about nonMarkovian models when the labels refer to compartments and sojourn time distributions are not exponential, while calling the labels ‘states’, even though, strictly speaking, they do not qualify as such.)

## 8 About the peak of the incidence curve

An epidemic curve has many features, such as

- the initial growth rate *ρ*;
- the height and timing of the peak;
- the final size.

For the first and last of these, it is well understood how they relate to the parameters of simple models that ignore heterogeneity. For instance, the final size is completely determined by *R*_0_, while *ρ* is a solution of the Euler-Lotka equation, cf. (4.5).

At the start of an outbreak, one may observe the initial growth rate and next use information about the generation interval to make inferences about *R*_0_, see [17, 18] and the references given there. Next one may choose the model parameters such that *ρ* and *R*_0_ of the model correspond to the estimated values.

The ongoing outbreak of Covid-19 generates much interest in peaks, largely because of concern that hospitals may be overwhelmed with patients, leading to healthcare breakdown. As far as we know, there is no analytical method to determine the height and timing of the peak from model parameters (except, perhaps, in the oversimplified *SIR* system of differential equations). So one has to rely on numerical calculations.

The key question addressed in this section is: how much is peak height influenced by model details? Here, we systematically compare the discrete-time *SEIR* model, described by (5.4) and corresponding to geometric distributions of the length of the latent and infectious period, to a model with deterministic, i.e., fixed, duration of these periods and constant infectiousness during the infectious period. Thus both types of model have three parameters. By restricting to *R*_0_ = 2.5 we fixed the infectiousness parameter in terms of the other two. We calibrated the models by making sure that *ρ* and the mean length of the latent period are the same, thus creating a one-to-one relationship between the two parameters of one type of model and the two parameters of the other type of model.

As initial condition we took a short history of decreasing fractions of susceptibles, reflecting an exponential increase in new cases at the rate *ρ*. We computed the peak value of the incidence for both types as a function of the two parameters and next their ratio. The results are depicted in Figure 2.

**Figure 2.**
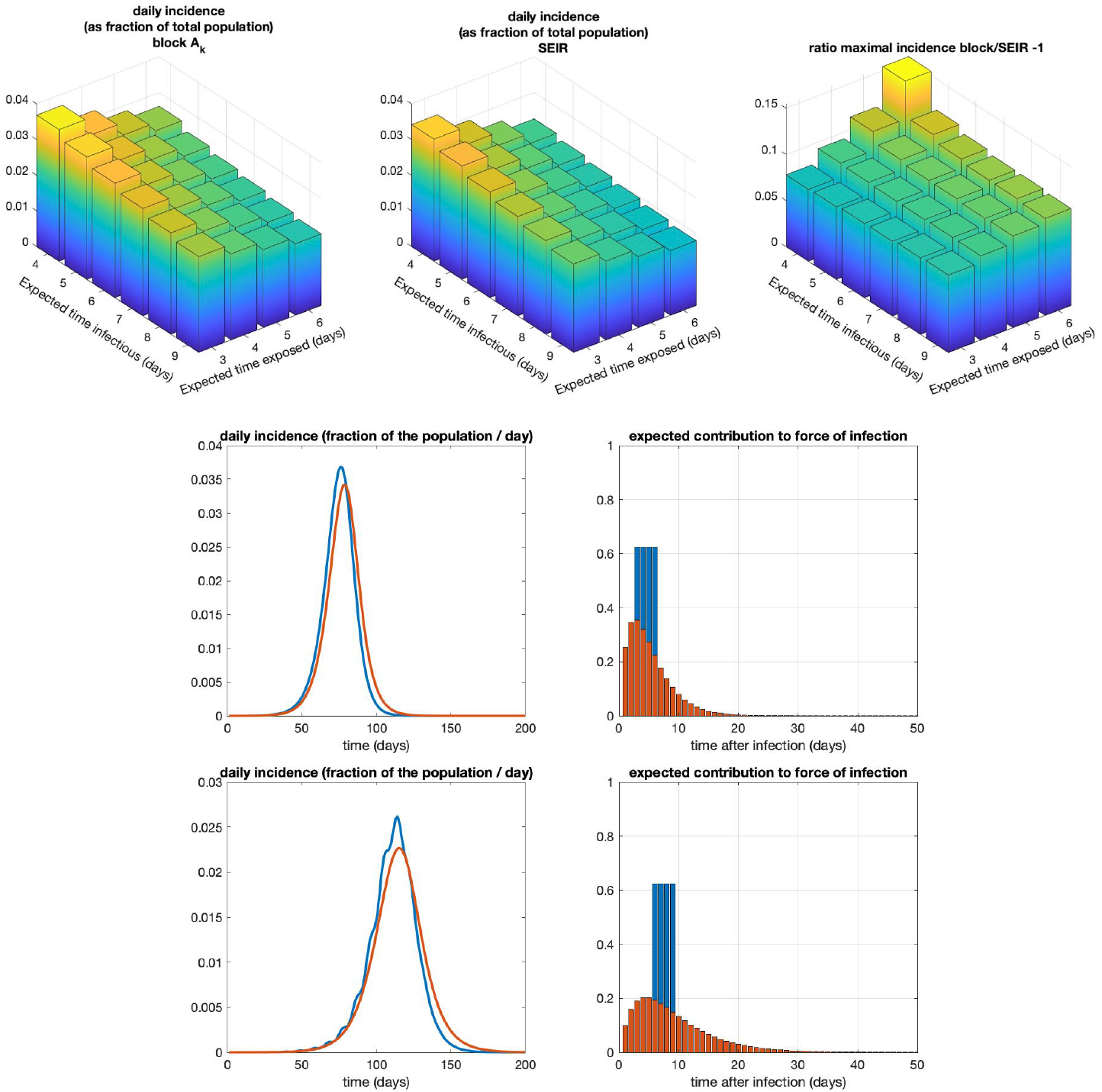
Comparing incidences between two types of models. In the block model, the lengths of the latent and infectious periods are deterministic (fixed for all individuals); in the *SEIR* model, the lengths of these periods are stochastic (independently exponentially distributed with identical parameters, respectively, 1*/T*_*E*_ and 1*/T*_*I*_). See Appendix for details. Top row: maximum incidence of the block model (with deterministic periods; left), *SEIR* model (with stochastic periods; middle) and the relative ratio between the two (i.e.,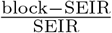; right), as a function of *T*_*E*_, the (actual resp. expected) time individuals are exposed and *T*_*I*_, the (actual resp. expected) time individuals are infectious. Models were compared after ensuring that they have the same *R*_0_ and initial speed *ρ*. Note that the incidence of the deterministic model always reaches a higher peak within the ranges of *T*_*E*_ and *T*_*I*_ considered, by about 8-15%, than the corresponding *SEIR* model. Middle and bottom rows: example simulations with the deterministic model (blue) and corresponding *SEIR* model (red) with the same *R*_0_ and *ρ*. The middle row corresponds to the parameters at which the ratio of peak heights is minimal, (*T*_*E*_, *T*_*I*_) = (3, 4); the bottom row to when this ratio is maximal, (*T*_*E*_, *T*_*I*_) = (6, 4). One can clearly see that the incidence grows initially at the same rate.

The main conclusions are:

- deterministic periods lead to higher peaks than geometrically distributed periods;
- this is most prominent when the latent period is large and the infectious period is small;
- the difference is, for reasonable parameter values, in the order of 10%.

(Note that, since compartmental models have fatter tails, they need, for given *R*_0_, to have an earlier peak of infectiousness in order to have the same *ρ*. This is clearly visible in Figures 2 and 4.)

After the first conclusion emerged, we aspired to find a somewhat mechanistic explanation. This led to the following observation. Roughly speaking, an outbreak reaches its peak when *S* is reduced to the level corresponding to *R*_0_ = 1. How many more cases there will be after the peak depends largely on the number of individuals that are, or are on the way of becoming, infectious at the time the peak is reached. (In [6, Section 1.3.2] it is explained how the overshoot phenomenon corresponding to a large stock of recently infected individuals at the time of reaching the peak, causes the final size, as fraction of the population, to increase when *R*_0_ increases.) For compartmental models, there is a relatively fat tail in the distribution of the time until becoming ‘removed’, i.e., having no future infectiousness. So when comparing models with the same *R*_0_, and hence the same final size, we should expect that for compartmental models the reservoir of latent and infectious individuals is less big, at peak-time, than for models in which expected future infectiousness reduces to zero after finite time. This is illustrated in Figure 3. And as reservoir size correlates with peak size, we should expect lower peaks for compartmental models, exactly as found in our numerical results.

**Figure 3.**
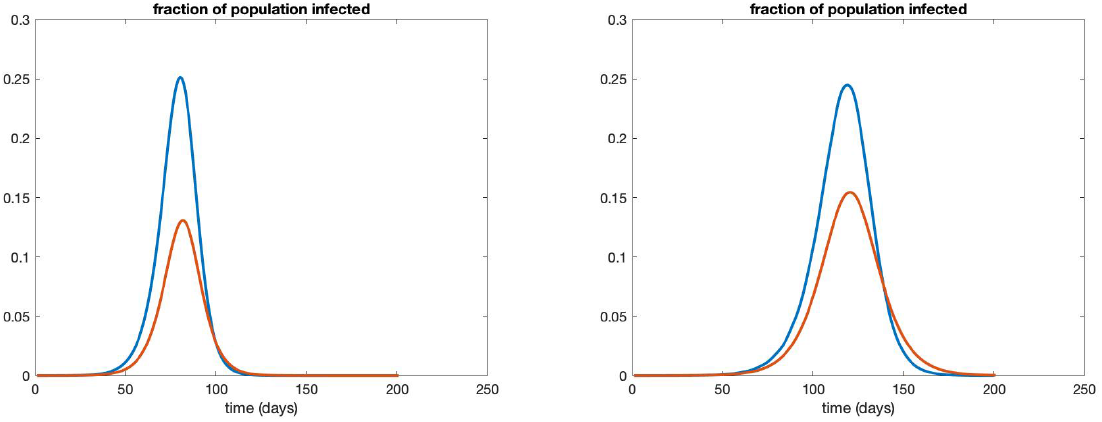
The fraction of the population that is either latently infected or infectious in the block model (blue) and the corresponding *SEIR* model. Left: (*T*_*E*_, *T*_*I*_) = (3, 4); right: (*T*_*E*_, *T*_*I*_) = (6, 4). The corresponding incidences can be found in Figure 2. Note that although the peak incidences are not so very different (see Fig. 2), there is a large difference in the fraction of infected-and-not-yet-removed individuals, due to the comparatively fatter tail in the expected future contribution to the force of infection in the *SEIR* model.

**Figure 4.**
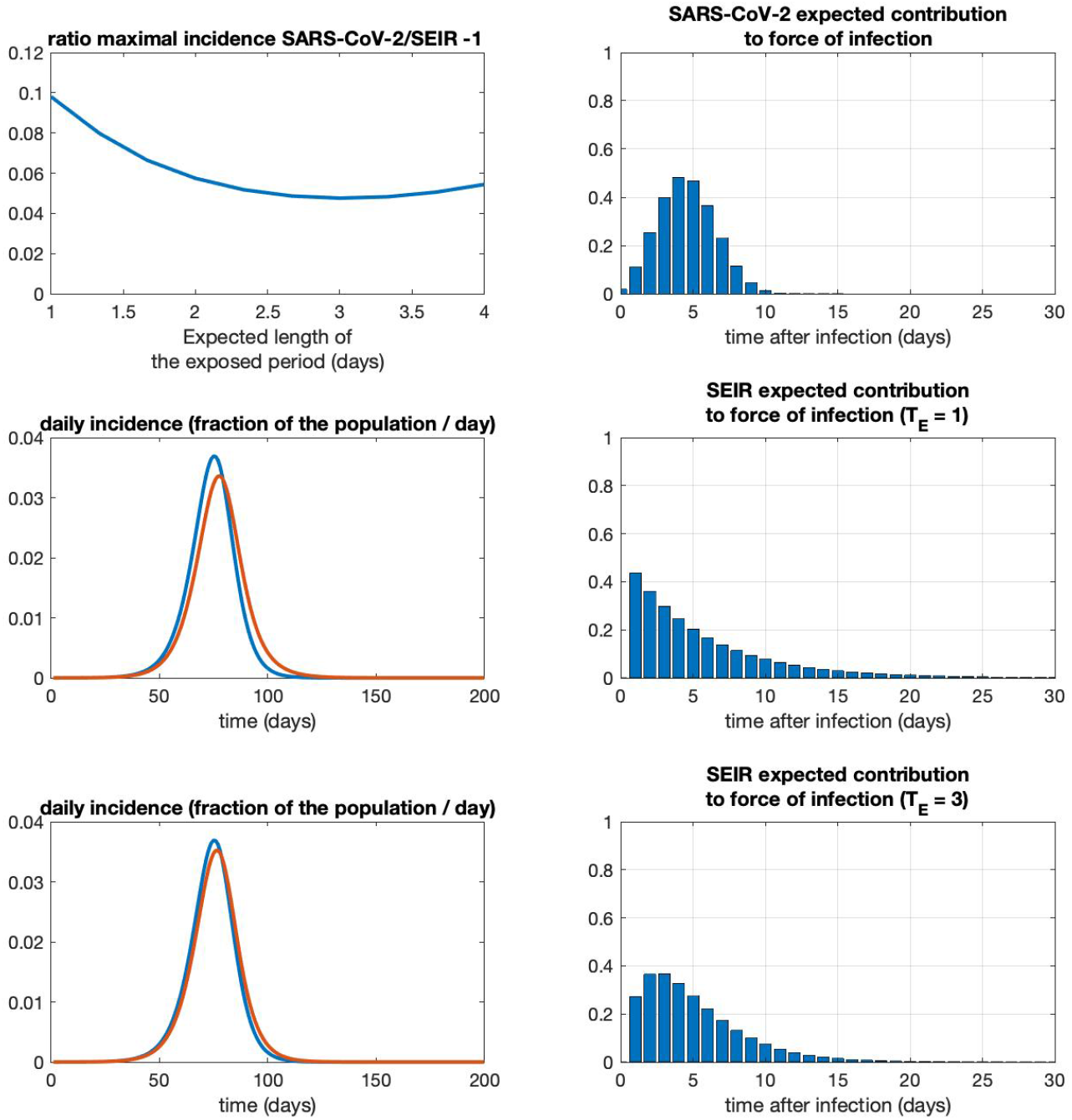
Comparing a model with parameters corresponding to the Weibull distribution derived from SARS-CoV-2 generation-interval data in [8] to the *SEIR* model with the same reproduction number *R*_0_ and the same initial growth rate *ρ*. Top row: left: relative ratio between the maximal incidences (*−* 1, as in Figure 2) as a function of *T*_*E*_, the expected length of the exposed period (note that the SARS-CoV-2 model does not depend on *T*_*E*_, see the Appendix). right: expected contribution to force of infection for the SARS-CoV-2 model. Middle row: left: incidence of the SARS-CoV-2 model (blue) and the *SEIR* model (red) with *T*_*E*_ = 1. At this *T*_*E*_, the relative difference in the peak incidences are maximal. right: the expected contribution of the *SEIR*-model for *T*_*E*_ = 1. Bottom row: same, but now for *T*_*E*_ = 3, at which the difference in peak incidence is minimal. For *T*_*E*_ *>* 4.12, the *SEIR*-model cannot be parameterized to have the same *R*_0_ and *ρ* as the SARS-CoV-2 model. For further details, see the Appendix.

Just to elucidate that the higher-peak-phenomenon matters in Covid-19 context, we chose, on the one hand, the parameters *A*_*k*_ as integrals over one day time-intervals of the Weibull generation-interval distribution as derived from data in [8] and, on the other hand, determined the one-parameter family of *SEIR* models that has both *R*_0_ and *ρ* equal to these quantities for the Weibull. The results of a comparison are presented in Figure 4. The peak heights differ 5 to 10%.

## 9 Conclusions

The success of the *SIR* and *SEIR* variants dwarfs the attention for the general Kermack-McKendrick model from 1927, even though, in principle, the latter has much on offer for a would-be modeller. We surmise that the reason is that the general model is formulated as a renewal (or, Volterra integral) equation and that for these unfamiliar equations there are no user friendly numerical tools available.

Here we introduced a discrete time version that has many advantages:

- the generality and flexibility is retained;
- computing the epidemic time course is super easy;
- the time step can be adjusted to the time interval between data points (e.g., one day or one week).

In Section 8 we showed that precise assumptions about the latent and infectious period matter for predicting the peak of the incidence curve, a quantity of interest from a public health perspective. So, we claim, the generality matters for practical issues and is not just an academic fancy. Of course in a practical context all kinds of heterogeneity (e.g., reflecting age) matter as well. These have been neglected here, but in the text book [6] and in [3] they have received ample attention in the continuous time setting, so it should not be too difficult to incorporate them in the discrete time framework as well.

Our hope is that our pragmatic reformulation leads to, well-deserved and long overdue [5], popularity of the true Kermack-McKendrick model.

## Data Availability

n/a

## Acknowledgements

It is a pleasure to thank Matthias Kreck for prompting one of us to formulate the discrete time Kermack-McKendrick model. In turn, it was the Covid-19 pandemic that sparked the interest of Matthias Kreck and Erhard Scholz, see [14, 15].

## 10 Appendix

### 10.1 Detailed description of numerical work displayed in Figure 2

In Figure 2 we numerically compare two types of models that have the same initial rate of increase *ρ* and basic reproduction number *R*_0_ but different expected contributions to the force of infection *A*_*k*_. Both models are defined by prescribing the values of the rescaled expected contribution to the force of infection, *Ã* _*k*_, where *k* is the number of days that have elapsed since becoming infected. Let us choose *T*_*E*_, the number of days an individual is exposed but not yet infectious, and *T*_*I*_, the number of days an individual is infectious. In the ‘block model’, in which these periods are assumed to be deterministic, so the same for all individuals, we set

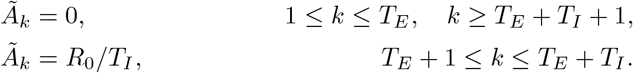

In this way, 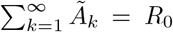. The initial exponential rate *ρ* with which the epidemic spreads is the solution to (4.5).

With *R*_0_, *ρ*, and *T*_*E*_, we can set up now the corresponding *SEIR* model. Using the explicit expression of the *A*_*k*_ for the *SEIR* model (5.5), we can determine the relation between *α, β* and *γ* on the one hand, and *ρ* on the other, using (4.5),

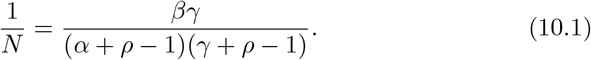

From the expression (4.3) for *R*_0_, we obtain the familiar expression 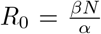. Substitution into (10.1) gives us

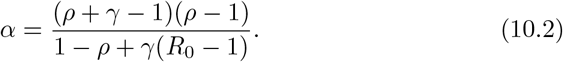

Lastly, the link between *γ* and *T*_*E*_ is simply 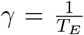. So in all, *T*_*E*_, *ρ* and *R*_0_ define the following parameters for the *SEIR* model,

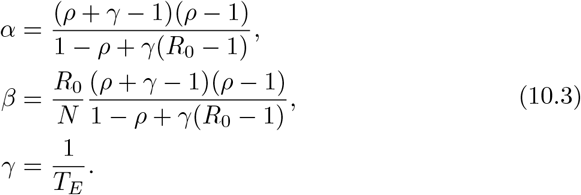

so that the *Ã* _*k*_ are given by

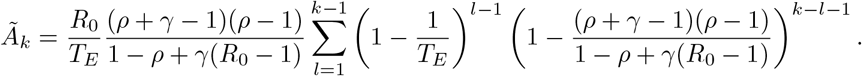

(The reader may verify that with this choice 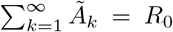, and that the characteristic equation (4.5) is satisfied exactly at *λ* = *ρ*, as required.)

The numerical simulations were carried out using (3.9), using an initial condition in which the epidemic has started increasing at rate *ρ* for six days:

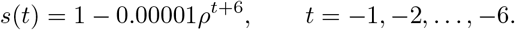

### 10.2 Figure 3: Comparing the *SEIR* model with a SARS-CoV-2 model

In [8], the generation interval distribution *g*(*τ*) is approximated by a Weibull distribution with shape parameter 2.826, and scale parameter 5.665. We discretized this,

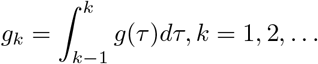

Then we set *Ã* _*k*_ = *R*_0_*g*_*k*_, *k* = 1, 2,…. The initial rate of increase is again estimated using 4.5, and gives *ρ* = 1.1919.

The corresponding *SEIR*-model is now given by using *α, β* and *γ* as in (10.3) and defining the *Ã* _*k*_ as before.

The initial condition is the same as in the previous illustration.

